# Identifying direct risk factors in UK Biobank with simultaneous Bayesian-frequentist model-averaged hypothesis testing using Doublethink

**DOI:** 10.1101/2024.01.01.24300687

**Authors:** Nicolas Arning, Helen R. Fryer, Daniel J. Wilson

## Abstract

Big data approaches to discovering non-genetic risk factors have lagged behind genome-wide association studies that routinely uncover novel genetic risk factors for diverse diseases. Instead, epidemiology typically focuses on candidate risk factors. Since modern biobanks contain thousands of potential risk factors, candidate approaches may introduce bias, inadequately control for multiple testing, and overlook important signals. Doublethink, a novel model-averaged hypothesis testing approach, offers a solution that simultaneously controls the Bayesian false discovery rate (FDR) and frequentist familywise error rate (FWER) while accounting for uncertainty in variable selection. Here we investigate direct risk factors for COVID-19 hospitalization from among 1,912 variables in 201,917 UK Biobank participants by implementing a Doublethink-based exposome-wide association study using Markov Chain Monte Carlo. Focusing on the 2020 outbreak, we find nine individual variables and six groups of variables exposome-wide significant at 9% FDR and 0.05% FWER. We identify significant direct effects among relatively overlooked risk factors including psychiatric disorders, dementia and prior infection, which we evaluate in relation to studies of other populations. We detect significant direct effects among some commonly reported risk factors like age, sex and obesity, but not others like diabetes, cardiovascular disease, hypertension, which may be mediated instead through variables representing general comorbidity. Doublethink produces interchangeable posterior odds and *p*-values for individual variables and arbitrary groups, facilitating flexible and powerful *post-hoc* hypothesis testing. We discuss the potential for impact and limitations of joint Bayesian-frequentist hypothesis testing, including the benefits of an agnostic exposome-wide approach to discovery.

**Significance:** Understanding what causes disease is key to improving its treatment and prevention. Large health studies like UK Biobank measure thousands of possible causes of disease. Traditionally, scientists have studied possible causes (like smoking or exercise) one-at-a-time, in depth. For greater perspective, we could study them altogether to test which have any effect. We recently introduced Doublethink, which combines the advantages of two major statistical approaches to testing. Here we use Doublethink to test 1,912 possible causes of COVID-19 hospitalization in UK Biobank. We found strong evidence for relatively overlooked causes: psychiatric conditions, dementia and previous infections. Findings from other health studies support these causes, highlighting the need to re-evaluate them and showing how our approach can reveal valuable insights.

## Introduction

The big data era has seen the advent of biobank-scale scans for genetic determinants of diverse health outcomes in cohorts like UK Biobank (1, 2). But similar data-driven identification of non-genetic determinants, termed risk factors, has not become commonplace. Instead, current epidemiology typically reports on candidate risk factors. Studies aim to address the question: What is the total effect of a variable on the outcome? Is it non-zero? For instance, more than 100 published studies have analysed dozens of candidate risk factors for COVID-19 outcomes in UK Biobank (Table S1).

Synthesizing these findings is difficult because: (i) Other, more important, risk factors that were not analysed may exist among the thousands measured. (ii) It is unclear how to appropriately limit false positives caused by multiple testing. (iii) The processes of selecting candidate risk factors and deciding to publish are vulnerable to bias. The experience of candidate gene studies, largely superseded by genome-wide association studies (GWAS), raises further questions about strength of evidence and reproducibility in candidate risk factor studies (3, 4, 5).

A major complication for systematic studies of non-genetic risk factors, compared to GWAS, is the problem of mediation (6). Mediation occurs when the total effect of a variable (e.g. age) on an outcome (e.g. COVID-19 severity) is wholly or partially caused through another variable (e.g. prior pneumonia). This conceptually divides the total effect into direct and indirect effects. Mediation is ignored in GWAS because genetic variables are coinherited at conception; they cannot generally cause one another. So the question in GWAS is effectively: What is the direct effect of a variable on the outcome? Is it non-zero? Artefactual signals generated by confounding are instead the major concern. Controlling for other variables helps avoid confounding (7), but controlling for mediating variables alters the scientific question by shifting attention from total to direct effects. Unfortunately, direct effects can differ in direction and magnitude to total effects, a source of bias known as the Table 2 fallacy (8). Further pitfalls include reverse causation and collider bias (9).

Nevertheless, the demand for GWAS-inspired exposome-wide association studies (10) presents an opportunity, which has been partly filled by machine learning (11, 12). Machine learning offers a data-driven agnostic approach. A major advantage is its ability to analyse high dimensional data with minimal intervention, even in the presence of collinearity and widespread correlation between variables. But the question in machine learning is different: What is the contribution of a variable to predicting the outcome? Usually there is no formal test. More importantly, a variable can be valuable for prediction due to confounding (13). Machine learning is therefore problematic for risk factor identification. Other concerns have been raised with artificial intelligence approaches in healthcare, particularly in terms of often difficult-to-achieve interpretability and equity (14).

Bayesian methodology offers a solution to the question of identifying direct effects in biobank-scale data while controlling for confounding (15). An important advantage is the ability to account for uncertainty in model choice by averaging over the inclusion or exclusion of other variables when estimating or testing the direct effect of each variable. This uncertainty can strongly influence conclusions. The question is therefore: What is the explanatory value of each variable, over and above all the other variables? Is it non-zero? With a careful approach to feature engineering to mitigate issues around mediation, reverse causality and collider bias, and with independent replication of discoveries, Bayesian model averaging (BMA) offers a powerful approach. But Bayesian approaches are seldom used in current epidemiology: none of 127 published studies of risk factors for COVID-19 outcomes in UK Biobank was Bayesian (Table S1). This might be explained by several issues, including lack of familiarity among researchers, high computational requirements, and difficulties specifying prior distributions (16). Many practitioners worry about the role of the prior in Bayesian hypothesis testing, which can lead different researchers to different conclusions from the same data (17).

Doublethink (18) is a new approach that aims to address these concerns by facilitating joint Bayesian-frequentist model-averaged hypothesis testing while simultaneously controlling the Bayesian false discovery rate (FDR) and the frequentist familywise error rate (FWER). By implementing Doublethink using a Markov Chain Monte Carlo (MCMC) algorithm, we are able to test for direct risk factors among thousands of individual variables, and arbitrary groups of those variables, while accounting for uncertainty in variable selection. We apply Doublethink to investigate direct risk factors for COVID-19 hospitalization in UK Biobank among 1,912 variables in 201,917 participants, and we compare our results to the literature. Our framework provides a highly capable model-averaging approach that can be applied to the systematic evaluation of direct risk factors in biobank-scale resources.

### Theory

We consider a general regression setting in which there are *n* observed outcomes *y*_1_ … *y_n_* and *ν* variables (features) with regression coefficients *β*_1_… *β_ν_*. The aim is to identify which variables directly influence the outcome, i.e. which of the regression coefficients are non-zero. In total there are 2*^ν^*hypotheses, *ω****_v_***, which we index with vector ***v***. The *j*th element of ***v*** indicates whether we are testing that variable *j* is zero (*v_j_* = 0; *β_j_* = 0) or not testing it (*v_j_* = 1; *β_j_* = 0 or *β_j_* ≠ 0).

In parallel, we define 2*^ν^* models with non-overlapping parameter spaces Θ**_s_**, indexed by vector ***s***, the *j*th element of which indicates whether variable *j* is zero (*s_j_* = 0; *β_j_* = 0) or non-zero (*s_j_* = 1; *β_j_* ≠ 0). Each null hypothesis *ω****_v_*** is compatible with one or more models Θ***_s_***, indexed by the set

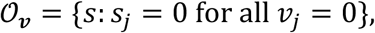

and incompatible with all the other models, indexed by the complementary set *A_v_*. In the Bayesian setting, we reject null hypothesis *ω****_v_*** if the posterior odds of models in *A_v_* versus *O_v_*,

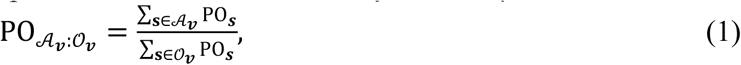

exceed some threshold *τ*. Here PO_*_ represents the posterior odds of model ***s*** versus the grand null model **0**. The Bayesian false discovery rate (FDR), both local and global (19), is then controlled at or below 1/(1 + τ), contingent on the prior.

Fryer, Arning and Wilson (18) showed that Bayesian hypothesis tests, like the above, inherently control the frequentist familywise error rate (FWER) in the strong sense because they are closed testing procedures (20). The FWER can be quantified, subject to further assumptions. Johnson (21, 22) developed an approach in which the posterior odds of model ***s*** versus model **0** can be approximated as

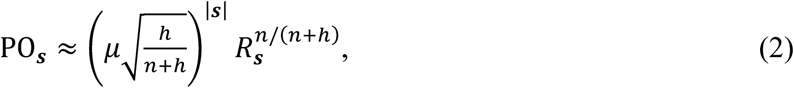

where *R_s_* represents the ratio of maximized likelihoods under model ***s*** versus model **0**, |***s***| gives the difference in the number of parameters to be estimated under model ***s*** versus model **0**, ; represents the prior odds that a regression coefficient is non-zero, *n* is the sample size, and *h* represents the precision of the prior on the non-zero regression coefficients. The approximation is based on assumptions including a large sample size, independent observations and the following prior:

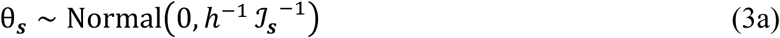

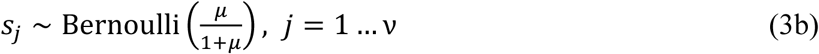

Here *θ****_s_*** represents the unconstrained parameters in model ***s*** (the *β_j_* for which *s_j_* = 1, and any nuisance parameters), and ℐ_*_ is the per-observation Fisher information matrix for model ***s***, evaluated at *θ****_s_*** = **0**. Fisher’s information matrix has been used widely in the definition of reference priors (e.g. 23, 24), and to generate concordance between Bayesian and frequentist point and interval estimates (see Table 1 of 18). Johnson’s approach converges on the Bayesian information criterion (BIC), which has been shown to reasonably approximate a wide range of posterior odds when *n* is large (25, 26). The strength of Johnson’s approach is the ability, for a pair of nested models, to interconvert posterior odds and *p*-values based on the likelihood ratio test (27).

**Table 1.** Doublethink allows the interconversion of model-averaged posterior odds and p-values for groups of variables, defined here a priori using variable correlation. The most significant groups are shown, alongside details of constituent variables. The most significant individual, ungrouped, variables are also shown. PP: posterior probability. p^*^: adjusted p-value. Dashes (-) indicate p^*^ > 0.02.

|  | Group<br>PP<br>(%) | Group<br>-log<sub>10</sub><br>p* | Variable | PP<br>(%) | -log<sub>10</sub><br>p* | Direct<br>effect<br>when<br>included | Standard<br>error<br>when<br>included |
| --- | --- | --- | --- | --- | --- | --- | --- |
| 1 | 100 | >5.95 | 34 Year of birth (years) | 40.8 | 2.05 | -0.03 | 0.00 |
|  |  |  | 21003 Age when attended assessment centre (years) | 30.0 | 1.82 | 0.03 | 0.10 |
|  |  |  | 21022 Age at recruitment (years) | 29.2 | 1.80 | 0.03 | 0.10 |
| 2 | 100 | >5.95 | 26413 Health score (England) | 77.7 | 2.83 | 0.26 | 0.03 |
|  |  |  | 26412 Employment score (England) | 10.9 | - | 2.79 | 0.37 |
|  |  |  | 26410 Index of Multiple Deprivation (England) | 9.9 | - | 0.01 | 0.00 |
|  |  |  | 189 Townsend deprivation index at recruitment | 2.7 | - | 0.05 | 0.02 |
|  |  |  | 26414 Education score (England) | 0.3 | - | 0.01 | 0.00 |
|  |  |  | 26411 Income score (England) | 0.3 | - | 1.70 | 0.34 |
|  |  |  | 26417 Living environment score (England) | 0.0 | - | 0.00 | 0.00 |
|  |  |  | 26416 Crime score (England) | 0.0 | - | 0.01 | 0.04 |
| 3 | 100 | 5.95 | 31 Sex : 0 : Female | 50.1 | 2.24 | -0.49 | 0.11 |
|  |  |  | 31 Sex : 1 : Male | 49.9 | 2.23 | 0.49 | 0.11 |
| 4 | 100 | 5.80 | 41214 Carer support indicators : 1 : Yes | 100 | 5.78 | 0.56 | 0.08 |
|  |  |  | 54 UK Biobank assessment centre : 11006 : Stoke | 0.0 | - | 0.24 | 0.21 |
| 5 | 99.9 | 5.70 | 6138 Qualifications : 3 : O levels/GCSEs or equivalent | 96.7 | 3.82 | -0.29 | 0.05 |
|  |  |  | 6138 Qualifications : 1 : College or University degree | 96.5 | 3.80 | -0.34 | 0.06 |
|  |  |  | 6138 Qualifications : 2 : A levels/AS levels or equivalent | 0.1 | - | -0.29 | 0.09 |
| - | 99.9 | 5.44 | Z86.4 Personal history of psychoactive substance abuse | 99.9 | 5.44 | 0.37 | 0.06 |
| 6 | 99.7 | 5.01 | F03 Unspecified dementia | 99.7 | 5.00 | 0.94 | 0.15 |
|  |  |  | G30.9 Alzheimer's disease, unspecified | 0.0 | - | 0.57 | 0.23 |
|  |  |  | F00.9 Dementia in Alzheimer's disease, unspecified | 0.0 | - | 0.59 | 0.26 |
| 7 | 99.7 | 4.89 | 137 Number of treatments/medications taken | 99.7 | 4.89 | 0.06 | 0.01 |
|  |  |  | 135 Number of self-reported non-cancer illnesses | 0.0 | - | 0.02 | 0.01 |
| - | 99.6 | 4.82 | J22 Unspecified acute lower respiratory infection | 99.6 | 4.82 | 0.51 | 0.09 |
| 8 | 99.5 | 4.71 | Z50.1 Other physical therapy | 79.9 | 2.89 | 0.53 | 0.09 |
|  |  |  | Z50.7 Occupational therapy and vocational rehabilitation, not elsewhere classified | 19.6 | - | 0.61 | 0.11 |
|  |  |  | Z50.5 Speech therapy | 0.0 | - | 0.34 | 0.22 |
| - | 99.2 | 4.47 | R29.6 Tendency to fall, not elsewhere classified | 99.2 | 4.47 | 0.63 | 0.11 |
| 9 | 98.2 | 4.10 | 41218 History of psychiatric care on admission : 8 : Not applicable | 98.2 | 4.10 | 0.49 | 0.09 |
|  |  |  | 41214 Carer support indicators : 99 : Not known | 0.0 | - | 0.03 | 0.19 |
| 10 | 89.0 | 3.22 | 48 Waist circumference (cm) | 89.0 | 3.22 | 0.02 | 0.00 |
|  |  |  | 21002 Weight (Kg) | 0.0 | - | 0.01 | 0.00 |
|  |  |  | 23098 Weight (Kg) | 0.0 | - | 0.00 | 0.00 |
| - | 84.1 | 3.02 | J18.1 Lobar pneumonia, unspecified | 84.1 | 3.02 | 0.49 | 0.09 |
| 11 | 80.0 | 2.89 | 21000 Ethnic background : 1001 : British | 70.2 | 2.64 | -0.40 | 0.08 |
|  |  |  | 1647 Country of birth (UK/elsewhere) : 6 : Elsewhere | 9.8 | - | 0.42 | 0.09 |
|  |  |  | 1647 Country of birth (UK/elsewhere) : 1 : England | 0.0 | - | -0.18 | 0.07 |
|  |  |  | 21000 Ethnic background : 1003 : Any other white background | 0.0 | - | -0.21 | 0.14 |
| - | 66.1 | 2.55 | K59.0 Constipation | 66.1 | 2.55 | 0.40 | 0.08 |
| 12 | 58.4 | 2.40 | N39.0 Urinary tract infection, site not specified | 58.4 | 2.40 | 0.40 | 0.08 |
|  |  |  | B96.2 Escherichia coli [E. coli] as the cause of diseases classified to other chapters | 0.0 | - | 0.32 | 0.12 |
| 13 | 40.1 | 2.04 | 3063 Forced expiratory volume in 1-second (FEV1) (litres) | 28.1 | 1.77 | -0.19 | 0.04 |
|  |  |  | 3062 Forced vital capacity (FVC) (litres) | 12.1 | - | -0.15 | 0.03 |
|  |  |  | 3064 Peak expiratory flow (PEF) (litres/min) | 0.0 | - | 0.00 | 0.00 |
| 14 | 40.1 | 2.04 | 2188 Long-standing illness, disability or infirmity : 0 : No | 21.6 | - | -0.25 | 0.06 |
|  |  |  | 2188 Long-standing illness, disability or infirmity : 1 : Yes | 18.5 | - | 0.25 | 0.06 |
| 15 | 38.3 | 2.00 | 24017 Nitrogen dioxide air pollution; 2006 (micro-g/m3) | 15.5 | - | 0.01 | 0.00 |
|  |  |  | 24018 Nitrogen dioxide air pollution; 2007 (micro-g/m3) | 15.1 | - | 0.01 | 0.00 |
|  |  |  | 24016 Nitrogen dioxide air pollution; 2005 (micro-g/m3) | 7.7 | - | 0.01 | 0.00 |
| - | 31.5 | 1.86 | L97 Ulcer of lower limb, not elsewhere classified | 31.5 | 1.86 | 0.69 | 0.14 |
| - | 25.5 | 1.71 | N18.9 Chronic renal failure, unspecified | 25.5 | 1.71 | 0.48 | 0.10 |

Doublethink (18) extends this approach to the multiple testing setting, in which there is model uncertainty. Using the theory of heavy-tailed random variables (28–31) and asymptotic likelihood theory (e.g. 32), we showed that rejecting all null hypotheses *ω****_v_*** for which 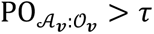 controls the FWER in the strong sense at or below level

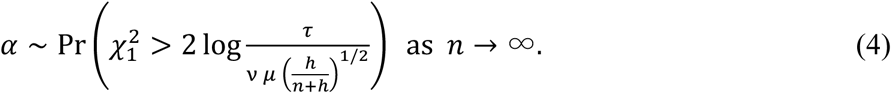

The Bayesian procedure is equivalent to rejecting the null hypothesis *ω***_v_** when an asymptotic *p*-value, adjusted for multiple testing,

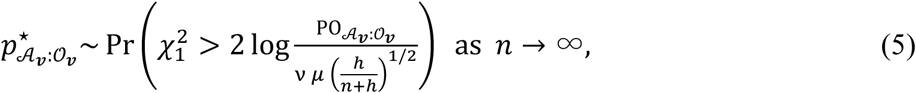

is smaller than threshold *α*. In general, the convergence in these asymptotic results is pointwise.

An equivalent interpretation of the results is that the test statistic 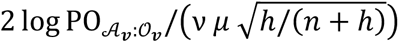, which follows a chi-squared distribution with one degree of freedom when large, represents a model-averaged deviance. Doublethink *p*-values cannot be trivially rescaled by the prior parameters *μ* and *h* because (i) the null distribution of the model-averaged deviance does not depend on them and (ii) the realized value depends on them only through weights. Therefore *μ* and *h* influence the power of the test, but not its large-sample theoretical distribution under the null hypothesis. This makes model-averaged hypothesis testing a workable frequentist method by facilitating a prior-agnostic approach to quantifying Bayesian significance thresholds in terms of frequentist FWER, for large samples. For further details, simulations, limitations of the approach such as inflation of the tests due to highly correlated variables, and mitigations such as testing groups of highly correlated variables, see (18).

## Methods

In this study, we developed a Monte Carlo Markov Chain algorithm (33) in R (34) and Python (35) that implements the Doublethink approach for thousands of variables, in order to identify direct risk factors for COVID-19 hospitalization in UK Biobank. We followed the COVID-19 Host Genetics Initiative definition of COVID-19 hospitalization, as applied to UK Biobank.

### Outcomes

Cases were identified from Public Health England’s Second Generation Surveillance System (SGSS), the National Health Service’s Hospital Episode Statistics (HES) and the National Health Service’s death registry between January and December 2020 as PCR positive for SARS-CoV-2 in SGSS, and hospitalized with International Classification of Diseases, Tenth Revision (ICD-10) diagnosis code U07.1 or U07.2 in HES. Participants not identified as cases were considered controls. We excluded participants that died before 2020, non-England residents determined by assessment centre, and those that withdrew before the analysis. The total number of controls was down-sampled to 200,000 to speed computation. The total number of cases was 1,917.

### Variables

We considered data fields approved for UK Biobank project 53100 ‘Microbiology, disease and genetics’, across the categories Population characteristics, Assessment centre, Biological samples, Online follow-up, Additional exposures and Health-related outcomes. We excluded Compound, Date, Text and Time variables, and variables concerning genetics and sampling processes. For repeated measures, we took the first instance. We excluded factors exceeding 50 levels, except self-reported illnesses, and variables missing in more than 15% of participants. Special values, including negative factor levels, were treated as missing. We imputed missing continuous and integer covariates taking the mean of non-missing values. Missing factor levels were treated as a separate level and excluded. We created binary variables for all levels of every factor observed with frequency above 0.2%. We created a binary variable for every ICD-10 code with frequency above 0.2% recorded before 2020 in HES. Overall, we analysed 184 covariates, binary variables encoding 865 levels across 193 factors, and 863 ICD-10 admission codes, a total of 1,912 variables (Supplementary Table S2).

### Model

We fitted the data separately for each outcome via a logistic regression model implemented in R using the glm function, assuming an additive linear predictor with an intercept term. We assumed the prior odds of variable inclusion were *μ* = 0.0053, independently for the *ν* = 1,912 variables, implying a prior expectation of 10 variables in the model. We assumed a unit information prior (*h* = 1) for the regression coefficients (26). We disallowed the inclusion of collinear variables by defining a zero likelihood.

### Implementation

In contrast to the Gibbs sampling approach of (36), we implemented a Metropolis-Hasting Markov Chain Monte Carlo (MCMC) sampler over the variable inclusion vector **s**. We ran 100 chains with 25,000 iterations of burn-in and 75,000 iterations of sampling. The average run-time per chain was 35 hours. Chains were initialized using a furthest neighbor algorithm to avoid including correlated variables. For initialization, we clustered variables into 200 groups with the scikit-learn-extra KMedoids algorithm, using rank correlation distance. Each chain was initialized with the medoid of one group, before adding nine more variables iteratively from the next-least correlated variables. Three Metropolis Hastings moves were implemented that respectively added, removed, or swapped pairs of variables with relative proposal probabilities 9:9:2. Variables were swapped preferentially for those with high squared correlation. We simulated regression coefficients directly from conditional Normal distributions by post-processing the MCMC iterations. We calculated posterior odds and parameter estimates by combining chains, computing Monte Carlo standard errors across independent chains.

### Grouping variables

We were able to perform valid *post-hoc* variable grouping while controlling the FDR and FWER, which was useful since correlated variables reduce one-another’s individual posterior inclusion probabilities. Moreover, tests of null hypotheses involving some but not all members of a cluster of highly correlated variables are liable to inflation (18). We clustered variables in two ways: (i) *A posteriori*, using the scikit-learn OPTICS algorithm (38) with distances defined by their posterior correlation in inclusion probabilities. This grouped the variables with the strongest negative correlations in inclusion probabilities. (ii) *A priori*, using the same algorithm applied to one minus the squared correlation between variables. We computed the posterior odds of including one or more of the variables in each group.

### *p*-value calculation

We used the chi-squared distribution to compute adjusted *p*-values using Equation 5. In the case of orthogonal variables with one degree-of-freedom, this is conservative for *p* < 0.02; see (18). Since the large sample size assumption implies interest in small significance thresholds, we report any *p*-value larger than 0.02 as n.s. (not significant) or ‘-’. Effectively, this makes Doublethink incompatible with any threshold exceeding *α* = 0.02. In practice, we did not explicitly set a threshold, instead reporting adjusted *p*-values alongside posterior odds.

### Literature review

We reviewed the variables included in published analyses of COVID-19 risk factors in UK Biobank using the query "UK Biobank" (Abstract) and "COVID" (Abstract) in www.webofscience.com on 19 September 2023. After excluding Review Articles and Editorial Material, this search returned 203 publications. We analysed a subset of 127 of these papers that quantified the effect of non-genetic risk factors on COVID-19 outcomes; this predominantly excluded papers reporting genetic risk factors, two-sample Mendelian randomization, and COVID-19 as an exposure for other outcomes (Supplementary Table S1). We manually categorized the variables analysed by these 127 papers into groups (Supplementary Table S3). We summarized the frequency with which each category of variable was included in the published analysis or abstract.

## Results

We aimed to identify risk factors that directly influenced COVID-19 hospitalization in UK Biobank participants to understand the underlying processes. We used model-averaged hypothesis tests to account for uncertainty in variable selection and deplete for potential confounders. We assumed the relevant variables were measured in UK Biobank. We aimed to limit the impact of collider bias by focusing on exposure variables measured before 2020, and by comparing cases to the rest of the biobank. This compounded the case definition with any selection biases in the sampling process, for example access to testing, which may affect interpretation (9). We focused on risk factors for hospitalization with COVID-19, because there were more cases than critical illness, and less obvious selection bias than infection, since testing was more widely available in hospitals.

### Doublethink facilitates joint Bayesian-frequentist model-averaged hypothesis tests

Figure 1 shows a Manhattan plot displaying the evidence that each of the 1,912 individual variables (circles) directly affected the risk of COVID-19 hospitalization in UK Biobank, averaged over uncertainty in the effect of all other variables. Points are plotted against both the log_10_ posterior odds (left side) and the −log_10_ adjusted *p*-value from Equation 5 (right side). This interconversion allows a Bayesian or frequentist approach to evaluating the strength of evidence.

**Figure 1.**
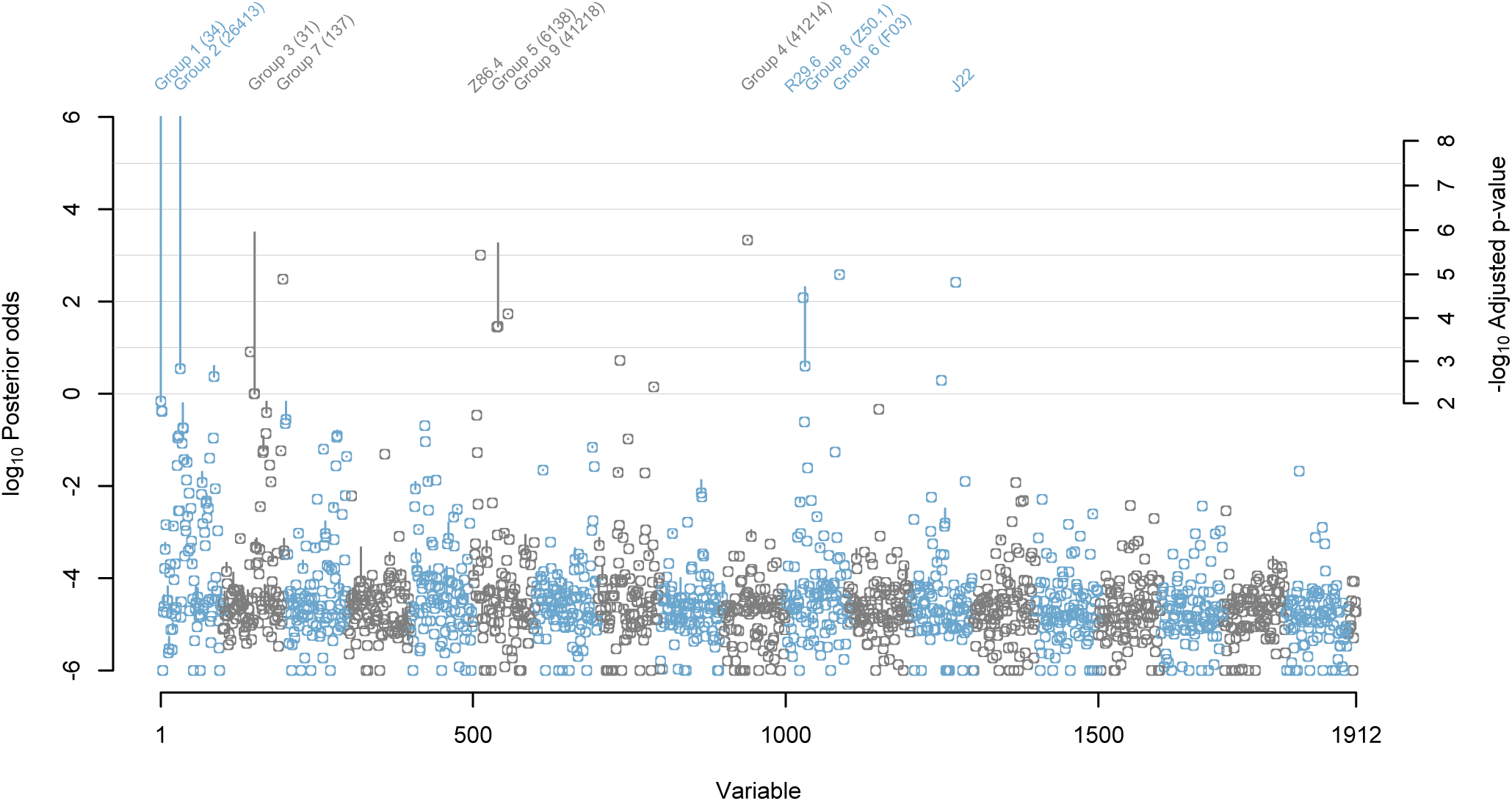
Individual variables (circles) and pre-defined groups of variables (vertical lines) with the strongest evidence of direct effects on the risk of COVID-19 hospitalization in UK Biobank. Evidence was quantified simultaneously by log_10_ posterior odds (left axis) and -log_10_ adjusted p-value (right axis) using Doublethink. Points and lines are coloured for legibility. Variables were ordered horizontally using OPTICS (38). Vertical lines show the boost in significance (if any) from the most significant individual variable per group to the significance of the whole group. Significance was truncated to log_10_ posterior odds between -6 and 6. Individual variables and groups significant at log_10_ posterior odds ≥ 1 are labelled (with the most significant variable per group in parentheses). Individual variables are named by UK Biobank field ID or, when prefixed by a letter, ICD-10 code. Refer to Table 1 for full names.

Comparison of the two vertical axis scales shows that in the Doublethink model, the model-averaged posterior odds and adjusted *p*-values are approximately linearly related, for small *p*-values. Significant variables are identified by applying a threshold to either the posterior odds or the adjusted *p*-value; this simultaneously controls the FDR (subject to the assumed prior) and the FWER (subject to the asymptotic approximation). For example, a Bayesian threshold of *τ* = 10 would control the FDR at 1/(1 + *τ*) = 0.091 and the FWER at *α* = 10^-3.3^ = 0.00047. The latter is much smaller than the conventional threshold of 0.05 because of the large sample size.

At a significance threshold of *τ* = 10 and *α* = 10^-3.3^, nine variables were identified as individually exposome-wide significant (Table 1). The interpretation is that significant variables, such as the ICD-10 codes `F03 Unspecified dementia` and `J22 Unspecified acute lower respiratory infection`, directly affect risk of COVID-19 hospitalization, even after controlling for the effects of all other measured variables. This differs from the common practice of testing the significance of a variable in the context of a single model that controls for a limited set of other variables. Model averaging is important in biobank-scale data where correlation between variables is pervasive, and no single model has high posterior probability.

For several significant variables, the interpretation that they directly affect risk of COVID-19 would seem too literal, such as `137 Number of treatments/medications taken`, which is based on the recruitment interview, `41214 Carer support indicators : 1 : Yes`, which indicates a hospital record of past carer support, `R29.6 Tendency to fall, not elsewhere classified`, which indicates a history and future risk of falls, and `Z86.4 Personal history of psychoactive substance abuse`, which indicates a hospital record of past alcohol, tobacco or drug use. More plausibly, these variables represent or aggregate one or more (perhaps unmeasured) variables that directly affect risk of COVID-19, like aspects of general health or behaviour. The estimated direct effect of these proxies was to increase the risk of COVID-19 hospitalization in all cases (Table 1). In contrast, significant measures of educational attainment, `6138 Qualifications : 3 : O levels/GCSEs or equivalent`, and `6138 Qualifications : 1 : College or University degree`, had protective direct effects on risk of COVID-19 hospitalization.

The significance of some variables was, at first glance, unexpectedly low, such as the well-established risk factors `31 Sex : 1 : Male` (Posterior probability, *PP* = 49.9; *p*^*^ = 10^−2.23^; where posterior odds = *PP*/(1-*PP*)) and `34 Year of birth (years)` (*PP* = 40.8; *p*^*^ = 10^−2.05^; Table 1). This is explained by the inclusion in the data of the other very highly correlated variables `31 Sex : 0 : Female`, `21003 Age when attended assessment centre (years)` and `21022 Age at recruitment (years)`. Including variables that are correlated, whether strongly or weakly, often dilutes the significance of individual variables when testing for the existence of a direct effect, over and above all other variables. For age and sex, an obvious solution would be to exclude these correlated variables – it may seem absurd not to have done so. However, correlation is pervasive in biobank-scale data, it may not be obvious which variables to exclude, and the impact of excluding variables on the results is hard to anticipate. An alternative solution is to define groups of correlated variables and test whether one or more members of a group directly affect the outcome. A major strength of Doublethink is it allows arbitrary groups of variables to be tested in this way, while controlling the FDR and FWER.

### Testing the significance of groups of variables reveals more signals

Nine groups of variables defined *a priori* were significant at *τ* = 10 and *α* = 10^-3.3^; for four groups, none of the member variables were individually significant. In Figure 1, vertical lines illustrate the boost in the significance of groups of variables compared to their most significant member. The groups are numbered for cross-reference with Table 1. Reassuringly, the well-established risk factors age (Group 1; *PP* = 100; *p*^*^ < 10^−5.95^), indices of multiple deprivation (Group 2; *PP* = 100; *p*^*^ < 10^−5.95^), and sex (Group 3; *PP* = 100; *p*^*^ = 10^−5.95^) were significant despite containing no individually significant member variables. In these examples, testing groups of variables recovered signal that was diluted by the inclusion in the data of highly correlated variables.

Another group was significant, despite containing no individually significant variables, demonstrating the ability of combined tests to detect subtle signals. Group 8 (*PP* = 99.5, *p*^*^ = 10^−4.71^) comprised `Z50.1 Other physical therapy` (*PP* = 79.9, *p*^*^ = 10^−2.89^) and `Z50.7 Occupational therapy and vocational rehabilitation, not elsewhere classified` (*PP* = 19.6, *p*^*^ > 0.02). These indicators of rehabilitation might represent or aggregate aspects of convalescence less well captured by the other 1,917 variables analysed. The analysis suggests this history of convalescence directly increased the risk of COVID-19 hospitalization, after controlling for all other measured variables.

Testing groups is useful but defining them *a priori* is not necessarily the most effective method of discovering signals, because the groupings might not be relevant to the outcome under investigation. For example, Group 8 also included variable `Z50.5 Speech therapy`, which appeared to contribute nothing to the group’s overall significance (*PP* = 0.0, *p*^*^ > 0.02). Conversely, failure to group relevant variables together can cause signals to be overlooked, as we see next.

### Doublethink allows arbitrary groups to be tested

One of the advantages of the Doublethink approach is it motivates the testing of arbitrary groups of variables without inflating the FWER or FDR through a multiple testing ‘fishing expedition’. This is because the thresholds of all possible tests are pre-defined in the closed testing procedure. Therefore we were free to search for the most significant groups of variables. To this end, we grouped variables post-hoc whose posterior inclusion probabilities (*PP*s) were negatively correlated, because this suggests they ‘competed’ for inclusion in the model.

Figure 2 and Table 2 show that post-hoc grouping revealed signals that were weaker in pre-defined groupings, such as post-hoc Group D (*PP* = 99.5, *p*^*^ = 10^−4.75^), which captured aspects of obesity by combining the individual variables `48 Waist circumference (cm)` (*PP* = 89.0, *p*^*^ = 10^−3.22^), `21001 Body mass index (BMI) (Kg/m2)` (*PP* = 5.5, *p*^*^ > 0.02) and `23104 Body mass index (BMI) (Kg/m2)` (*PP* = 5.0, *p*^*^ > 0.02). In contrast, prior Group 10 (*PP* = 89.0, *p*^*^ = 10^−3.22^) had only combined `48 Waist circumference (cm)` with the non-significant variables `21002 Weight (Kg)` and `23098 Weight (Kg)` (Table 1).

**Figure 2.**
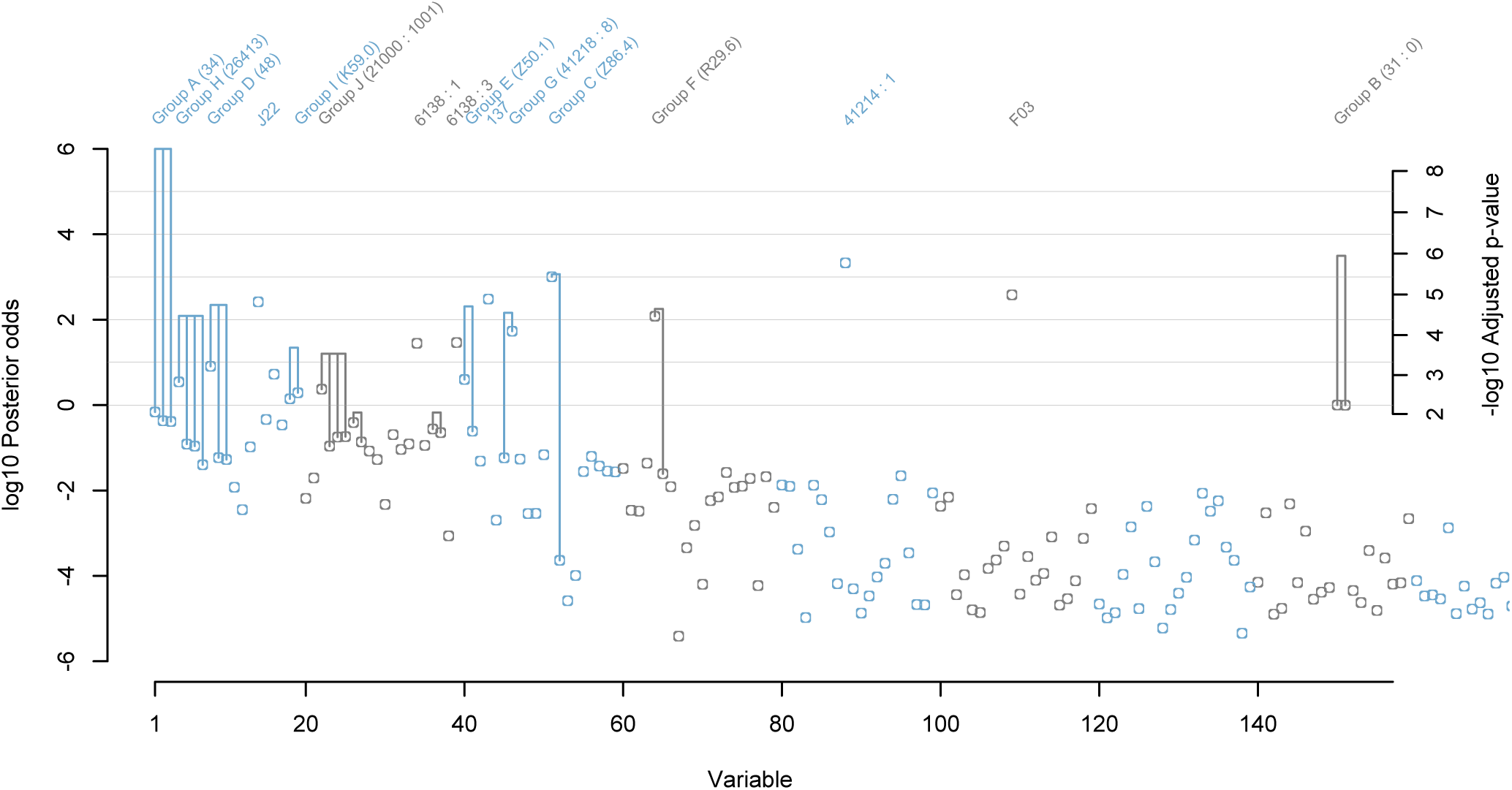
Individual variables (circles) and post-hoc groups of variables (vertical lines) with the strongest evidence of direct effects on the risk of COVID-19 hospitalization in UK Biobank. The horizontal axis is truncated to show only the most significant variables and groups. Vertical lines show the boost in significance (if any) from individual member variables to the significance of the whole group. See Figure 1 legend for further details.

**Table 2.** Doublethink allows arbitrary groups of variables to be assessed for significance while simultaneously controlling the FWER and FDR. Here groups were defined a posteriori by identifying variables whose PPs were negatively correlated. The most significant groups are shown, alongside details of constituent variables. The most significant individual, ungrouped, variables are also shown. PP: posterior probability. p^*^: adjusted p-value. Dashes (-) indicate p^*^ > 0.02.

|  | Group<br>PP<br>(%) | Group<br>-log<sub>10</sub> p* | Variable | PP (%) | -log<sub>10</sub><br>p* | Direct<br>effect<br>when<br>included | Standard<br>error<br>when<br>included |
| --- | --- | --- | --- | --- | --- | --- | --- |
| A | 100 | >5.95 | 34 Year of birth (years) | 40.8 | 2.05 | -0.03 | 0.00 |
|  |  |  | 21003 Age when attended assessment centre (years) | 30.0 | 1.82 | 0.03 | 0.10 |
|  |  |  | 21022 Age at recruitment (years) | 29.2 | 1.80 | 0.03 | 0.10 |
| B | 100 | 5.95 | 31 Sex : 0 : Female | 50.1 | 2.24 | -0.49 | 0.11 |
|  |  |  | 31 Sex : 1 : Male | 49.9 | 2.23 | 0.49 | 0.11 |
| - | 100 | 5.78 | 41214 Carer support indicators : 1 : Yes | 100.0 | 5.78 | 0.56 | 0.08 |
| C | 99.9 | 5.50 | Z86.4 Personal history of psychoactive substance abuse | 99.9 | 5.44 | 0.37 | 0.06 |
|  |  |  | 20116 Smoking status : 0 : Never | 0.0 | - | -0.17 | 0.07 |
| - | 99.7 | 5.00 | F03 Unspecified dementia | 99.7 | 5.00 | 0.94 | 0.15 |
| - | 99.7 | 4.89 | 137 Number of treatments/medications taken | 99.7 | 4.89 | 0.06 | 0.01 |
| - | 99.6 | 4.82 | J22 Unspecified acute lower respiratory infection | 99.6 | 4.82 | 0.51 | 0.09 |
| D | 99.5 | 4.75 | 48 Waist circumference (cm) | 89.0 | 3.22 | 0.02 | 0.00 |
|  |  |  | 21001 Body mass index (BMI) (Kg/m2) | 5.5 | - | 0.04 | 0.01 |
|  |  |  | 23104 Body mass index (BMI) (Kg/m2) | 5.0 | - | 0.04 | 0.01 |
| E | 99.5 | 4.71 | Z50.1 Other physical therapy | 79.9 | 2.89 | 0.53 | 0.09 |
|  |  |  | Z50.7 Occupational therapy and vocational rehabilitation, not elsewhere classified | 19.6 | - | 0.61 | 0.11 |
| F | 99.4 | 4.65 | R29.6 Tendency to fall, not elsewhere classified | 99.2 | 4.47 | 0.63 | 0.11 |
|  |  |  | W19.0 Home | 2.4 | - | 0.59 | 0.15 |
| G | 99.3 | 4.55 | 41218 History of psychiatric care on admission : 8 : Not applicable | 98.2 | 4.10 | 0.49 | 0.09 |
|  |  |  | I10 Essential (primary) hypertension | 5.5 | - | 0.24 | 0.06 |
| H | 99.2 | 4.48 | 26413 Health score (England) | 77.7 | 2.83 | 0.26 | 0.03 |
|  |  |  | 26412 Employment score (England) | 10.9 | - | 2.79 | 0.37 |
|  |  |  | 26410 Index of Multiple Deprivation (England) | 9.9 | - | 0.01 | 0.00 |
|  |  |  | 54 UK Biobank assessment centre : 11011 : Bristol | 3.9 | - | -0.47 | 0.14 |
| - | 96.7 | 3.82 | 6138 Qualifications : 3 : O levels/GCSEs or equivalent | 96.7 | 3.82 | -0.29 | 0.05 |
| - | 96.5 | 3.80 | 6138 Qualifications : 1 : College or University degree | 96.5 | 3.80 | -0.34 | 0.06 |
| I | 95.7 | 3.69 | K59.0 Constipation | 66.1 | 2.55 | 0.40 | 0.08 |
|  |  |  | N39.0 Urinary tract infection, site not specified | 58.4 | 2.40 | 0.40 | 0.08 |
| J | 94.1 | 3.54 | 21000 Ethnic background : 1001 : British | 70.2 | 2.64 | -0.40 | 0.08 |
|  |  |  | 24017 Nitrogen dioxide air pollution; 2006 (micro-g/m3) | 15.5 | - | 0.01 | 0.00 |
|  |  |  | 24018 Nitrogen dioxide air pollution; 2007 (micro-g/m3) | 15.1 | - | 0.01 | 0.00 |
|  |  |  | 1647 Country of birth (UK/elsewhere) : 6 : Elsewhere | 9.8 | - | 0.42 | 0.09 |
| - | 84.1 | 3.02 | J18.1 Lobar pneumonia, unspecified | 84.1 | 3.02 | 0.49 | 0.09 |
| K | 40.1 | 2.04 | 3063 Forced expiratory volume in 1-second (FEV1) (litres) | 28.1 | 1.77 | -0.19 | 0.04 |
|  |  |  | 3062 Forced vital capacity (FVC) (litres) | 12.1 | - | -0.15 | 0.03 |
| L | 40.1 | 2.04 | 2188 Long-standing illness, disability or infirmity : 0 : No | 21.6 | - | -0.25 | 0.06 |
|  |  |  | 2188 Long-standing illness, disability or infirmity : 1 : Yes | 18.5 | - | 0.25 | 0.06 |
| - | 31.5 | 1.86 | L97 Ulcer of lower limb, not elsewhere classified | 31.5 | 1.86 | 0.69 | 0.14 |
| - | 25.5 | 1.71 | N18.9 Chronic renal failure, unspecified | 25.5 | 1.71 | 0.48 | 0.10 |

Some post-hoc groups overlapped the pre-defined groups but dropped non-significant variables that did not contribute to the significance of the group. For example, Group H (*PP* = 99.2, *p*^*^ = 10^−4.48^), contained only the three most significant deprivation scores of the eight members of Group 2. Other groups revealed new connections between variables, such as Group I (*PP* = 95.7, *p*^*^ = 10^−3.69^), which combined the individually non-significant `K59.0 Constipation` (*PP* = 66.1, *p*^*^ = 10^−2.55^) and `N39.0 Urinary tract infection, site not specified` (*PP* = 58.4, *p*^*^ = 10^−2.40^). The former may cause the latter, but it is unclear how a history of these conditions could predispose to COVID-19 hospitalization.

The post-hoc grouping of `41218 History of psychiatric care on admission : 8 : Not applicable` with `I10 Essential (primary) hypertension` was at first glance surprising from the field descriptions (Group G: *PP* = 99.3, *p*^*^ = 10^−4.55^). However, the former variable indicates a history of non-psychiatric hospital care. This suggests it may act, in a manner interchangeable with `I10`, as a proxy for a history of underlying poor physical health. The direct effect of both variables was to increase the risk of COVID-19 hospitalization (Table 2).

The ability to quantify the evidence for groups of variables offers an alternative to approaches such as pre-analysis selection of representative candidate variables among groups of correlated variables. Doublethink permits all and any groups of variables to be tested while controlling the FDR and FWER. This presents new possibilities for identifying significant groups, and the identification of these groups may help with the interpretation of the role in the individual variables in the outcome.

### Comparison to the literature on COVID-19 outcomes in UK Biobank

Since early in the COVID-19 pandemic, before the discovery of effective treatments, there were intense research efforts to understand susceptibility to infection, disease and poor outcomes. Many studies focused on large established cohorts like UK Biobank that could rapidly link to data on SARS-CoV-2 testing (37), COVID-19 hospitalization (39) and mortality (40). Since then, many risk factors have been reported, including diabetes (41, 42, 43, 44), asthma (45, 46) and vitamin D (47, 48) as predisposing to worse outcomes. We compared our results to the literature on COVID-19 in UK Biobank to identify any differences to standard approaches and find new insights. At the time of analysis, we identified 127 comparable studies through Web of Science. We manually assigned the most common risk factors in published analyses of COVID-19 outcomes to larger categories for comparison to the variables and groups listed in Tables 1 and 2, which we assigned to the same list of categories.

Table 3 shows the most common categories of risk factors included in published analyses of COVID-19 outcomes in the 127 UK Biobank studies. Two summaries are shown: the percentage of papers and the percentage of abstracts in which each category of risk factors appeared. Alongside we show the evidence from our analysis, with values of *PP* < 50% (corresponding to *p*^*^ > 10^−2.20^) omitted, since the Bayesian interpretation is that this represents evidence against a direct effect of those risk factors. An important caveat is that many studies focused on the total (direct and indirect) effects of candidate risk factors, which our method does not quantify. Therefore, the discovery of a risk factor absent from the literature is more interesting than the non-discovery of (a possibly indirect) effect reported in the literature.

**Table 3.** Comparison of risk factors for COVID-19 outcomes in previous UK Biobank studies versus this study. The number of papers, out of 127, are shown. Categories were assigned manually from a literature review, and from Tables 1 and 2. When there were multiple matches in Tables 1 and 2, the maximum significance is given. PP: posterior probability (only values above 50% are shown). p^*^: adjusted p-value (only values below 10^−2.2^ are shown).

| Category | % Papers | % Abstracts | PP (%) | -log<sub>10</sub> p* |
| --- | --- | --- | --- | --- |
| Age | 90 | 11 | 100 | >5.95 |
| Sex | 84 | 14 | 100 | 5.95 |
| Obesity | 78 | 20 | 99.5 | 4.75 |
| Ethnicity | 78 | 16 | 80 | 2.89 |
| Socioeconomic status | 68 | 13 | 100 | >5.95 |
| Smoking | 66 | 6 | 99.9 | 5.5 |
| Diabetes | 63 | 10 |  |  |
| Cardiovascular disease | 59 | 12 |  |  |
| Hypertension | 54 | 11 |  |  |
| Lung disease | 46 | 6 | 99.6 | 4.82 |
| Alcohol intake | 35 | 3 | 99.9 | 5.5 |
| General comorbidity | 31 | 9 | 99.7 | 4.89 |
| Cancer | 29 | 1 |  |  |
| Kidney disease | 28 | 6 | 95.7 | 3.69 |
| Educational attainment | 28 | 4 | 99.9 | 5.7 |
| Asthma | 26 | 3 |  |  |
| Physical activity | 24 | 6 |  |  |
| Neurological disease | 21 | 3 |  |  |
| Liver disease | 19 | 2 |  |  |
| Inflammatory disease | 17 | 2 |  |  |
| Geographic region | 17 | 0 |  |  |
| Aging | 15 | 9 | 99.4 | 4.65 |
| Dementia | 15 | 2 | 99.7 | 5.01 |
| Employment | 13 | 3 |  |  |
| Immune disease | 12 | 2 |  |  |
| Diet | 11 | 6 |  |  |
| Depression | 11 | 4 |  |  |
| Infection | 10 | 3 | 99.6 | 4.82 |
| Arthritis | 10 | 3 |  |  |
| Other | 9 | 2 |  |  |
| Sleep disturbance | 9 | 6 |  |  |
| Psychiatric disorders | 9 | 5 | 100 | 5.8 |
| Mental health | 8 | 4 |  |  |
| Vitamin D | 8 | 4 |  |  |
| Lipid disorders | 7 | 2 |  |  |
| Pollution | 5 | 2 |  |  |
| Covid-19 related | 4 | 3 |  |  |
| Vaccination | 4 | 3 |  |  |
| Allergy | 4 | 1 |  |  |
| Haematological disease | 3 | 1 |  |  |
| Lifestyle | 3 | 2 |  |  |
| Gastrointestinal disease | 3 | 2 |  |  |
| Sex hormones | 2 | 2 |  |  |
| Periodontal disease | 2 | 2 |  |  |

Age, Sex, Obesity, Ethnicity, Socioeconomic status (including deprivation indices) and Smoking were included in 66-90% of published analyses, but mentioned in just 6-20% of abstracts. Our analysis supported direct effects of all these categories with *PP* ≥ 99.5% and *p*^*^ ≤ 10^−4.75^ except ethnicity (Supplementary Table S4, S5). Post-hoc Group J, which combined self-reported ethnicity and country of birth with geographic measures of pollution, had strong support (*PP* = 94.1, *p*^*^ = 10^−3.54^). However, pre-defined Group 11, which combined self-reported ethnicity and country of birth without pollution metrics, was not significant at *τ* = 10 and *α* = 10^-3.3^ (*PP* = 80.0, *p*^*^ = 10^−2.89^).

Other reasonably common categories of risk factor for which our analysis found evidence of direct effects included Lung disease, Alcohol intake, General comorbidity, Kidney disease and Educational attainment. Risk factors in these categories featured in 28-46% of published analyses and 4-9% of abstracts. Our analysis supported these categories with *PP* ≥ 95.7% and *p*^*^ ≤ 10^−3.69^.

Many categories of risk factors that appeared commonly in published analyses received no significant support for direct effects in our analysis. Diabetes, Cardiovascular disease and Hypertension were notable for inclusion in 54-63% of published analyses, and 10-12% of abstracts. No variables or groups of variables corresponding to these categories received support for direct effects in our analyses (*PP* < 50%, *p*^*^ > 10^−2.20^). However, no evidence of a direct effect does not imply no evidence of an effect. These common diseases contribute to a general decline of health, and it is possible that their effects were mediated through pathways better represented by variables or groups we categorised under General comorbidity, such as `137 Number of treatments/medications taken` and Group G (non-psychiatric hospital care/hypertension). Mediation is not the only explanation; the sparsity-favouring prior may have penalized the inclusion of direct effects of several variables in favour of an aggregate variable like `137 Number of treatments/medications taken` that captured the signal more parsimoniously.

Several notable categories of risk factor that we found to have significant direct effects were included infrequently in published analyses of COVID-19 outcomes in UK Biobank. Variables representing Psychiatric disorders, Dementia, Aging (over and above Age) and Infection were included in 9-15% of published analyses, and 2-9% of abstracts, whereas we found strong evidence of direct effects of variables we assigned to these categories (*PP* ≥ 99.4% and *p*^*^ ≤ 10^−4.65^), including `41214 Carer support indicators : 1 : Yes` (which we categorised under Psychiatric disorders), `F03 Unspecified dementia` (Dementia), `R29.6 Tendency to fall, not elsewhere classified` (Aging) and `J22 Unspecified acute lower respiratory infection` (Infection). These variables were significant even after accounting for all others, such as age and number of treatments/medications. Therefore a model-averaging big data approach that accounts for widespread correlations among variables and uncertainty in variable selection can bring useful perspective on our understanding of well-studied health outcomes like COVID-19 hospitalization in UK Biobank.

## Discussion

All exposome-wide significant direct effects we discovered had received some attention among the 127 UK Biobank studies in our literature review. However, we found several strongly significant signals in categories of variable overlooked by 85% of studies or more. Since our analysis investigated the same cohort, we drew on studies from other populations to assess the plausibility of these signals.

Psychiatric disorders were investigated by twelve out of 127 UK Biobank studies (9%), several of which reported an association with elevated risk of hospitalization and mortality (49–60). We found an exposome-wide significant signal that hospitalization risk was directly increased by a history of psychiatric hospital care, specifically among patients with a carer available at their usual place of residence. Supporting this, US studies found psychiatric diagnosis to be a risk factor for COVID-19 diagnosis (61, 62). In a matched case-control analysis of 3.5 million patients, diagnosis with a psychiatric disorder (psychotic disorder, mood disorder, anxiety disorder or insomnia) in the year before the outbreak increased the risk of COVID-19 diagnosis by 65% (61). Following infection, patients with recent diagnosis of mental disorders encompassing ADHD, bipolar disorder, depression and schizophrenia had 1.5-fold higher risk of hospitalization and 1.8-fold higher risk of mortality (62). A meta-analysis of 23 studies reported a two-fold increase in COVID-19 mortality risk in individuals with psychotic disorders (63). Proposed mechanisms include reduced adherence to social distancing, immune-inflammatory dysregulation in psychiatric conditions, and interactions with psychotropic medications (62, 63).

Dementia was investigated by 19/127 UK Biobank studies (15%), reporting increased risk of infection, hospitalization and mortality, with Alzheimer’s disease showing the highest risk of COVID-19 diagnosis and mortality (42, 53, 58, 59, 64–78). We found exposome-wide significant evidence that prior hospitalization with unspecified dementia directly increased the subsequent risk of COVID-19 hospitalization. In the US, analyses of electronic health records found that dementia increased the risk of COVID-19 diagnosis two-fold, with the strongest effect (3.2-fold) for vascular dementia (79), and increased the risk of mortality with COVID-19 by 1.3-fold (80). Proposed mechanisms for a causal effect of dementia on COVID-19 outcomes include compromised social distancing, particularly in care home settings, challenges maintaining personal hygiene, physical frailty, dementia-associated inflammation and immune dysregulation, and direct aggravating effects of viral infection on cardiovascular and respiratory brain function. Interestingly, imaging of 785 UK Biobank participants revealed brain tissue damage following SARS-CoV-2 infection (81), and the ACE2 receptor, by which SARS-CoV-2 invades human cells, is reportedly upregulated in the brains of Alzheimer’s patients (82).

Whereas age was investigated by 114/127 UK Biobank studies (90%), molecular and physical signs of aging were investigated by just 19/127 studies (15%). Phenotypic age acceleration (which estimates excess aging via blood biomarkers (83)), shorter leukocyte telomere length, physical frailty including falls, and slower walking pace were associated with worse COVID-19 outcomes (49, 50, 66, 72, 84–98). We found exposome-wide significant evidence that a prior hospital diagnosis of a tendency to fall directly increased the risk of COVID-19 hospitalization. Falls have been identified as an atypical presenting symptom of COVID-19 in some patients, and serve as a marker of underlying frailty (99). Frailty, frequently measured through a subjective clinical frailty score (100), has been reported as a risk factor for COVID-19 mortality in multiple countries, with three meta-analyses supporting the conclusion that frailty increases risk even after accounting for age (101–103).

Whereas lung disease, including prior pneumonia, was commonly investigated (58/127 UK Biobank studies; 46%), other markers of infection were investigated infrequently (13/127 studies; 10%). In those studies, elevated immune cell counts, infection-related biomarkers and virus antibody titres were associated with increased COVID-19 infection and severity (59, 66, 70, 76, 89, 104–110). We found exposome-wide significant evidence for an increased risk of COVID-19 hospitalization directly associated with prior hospital episodes of (i) unspecified acute lower respiratory infection and (ii) constipation or urinary tract infection. In other populations, prior pneumonia, lung disease and genetic susceptibility to lung disease have been identified as risk factors for COVID-19 (111–116). It is unclear why prior hospital episodes of constipation or urinary tract infection should increase the risk of subsequent COVID-19 hospitalization (117), although perhaps interestingly, ACE2 is reportedly expressed in kidney, bladder and intestine(118, 119). Pre-existing pathologies in these tissues might be exacerbated directly by the cell tropism of SARS-CoV-2 infection.

We did not find exposome-wide significant evidence for direct effects of some previously reported risk factors, like diabetes (41, 42, 43, 44), asthma (45, 46) and vitamin D (47, 48). This does not rule out indirect effects, but it highlights an important contrast with variables for which we did find significant evidence of direct effects, like the self-reported number of treatments/medications taken, prior hospital diagnosis of acute lower respiratory infection, and a history of medically-relevant psychoactive substance abuse (including alcohol and tobacco). There are several reasons we may not have detected direct risk factors. (i) The exposome-wide approach demands stringent multiple testing, reducing power. (ii) We employed a nominal FWER of *α* = 10^-3.3^, 100-fold more stringent than the conventional threshold 0.05. Our FWER threshold was interconverted from an FDR of 0.09 using Doublethink (18), but stringent FWER thresholds such as this improve replicability (120) and scaling the FWER with 1/√*n* ensures the consistency of hypothesis testing in large samples (121). (iii) The Doublethink framework incorporates a prior distribution with hyperparameters *μ* and *h* that affect inference. (iv) The Bayesian prior penalizes complex models, which may favor aggregate variables (like number of treatments/medications) over a set of individual risk factors (like diabetes and hypertension).

A definitive approach to detecting direct risk factors is unattainable, even for a systematic, agnostic scan, because direct effects are defined only relative to a fixed set of variables. A “direct” effect could be mediated through one or more downstream variables that were not measured. No method is free of subjectivity: we curated variables to uphold quality control, avoid reverse causation and avoid collider bias (e.g. by restricting analysis to pre-2020 exposures), and to fix outcome definitions (restricting attention to 2020 due to time-varying vaccine effects). We restricted analysis to 1,912 variables measured in 85% of more of the cohort. The methods chosen to impute missing values, handle repeat measures and encode factors further shaped our results.

Our work offers a new approach at a time when there are increasing calls for exposome-wide association studies (e.g. 10). Agnostic exposome-wide approaches to discovering new risk factors were rare among published studies of COVID-19 outcomes in UK Biobank (Table S1). A few studies applied machine learning and univariable scans (9, 12, 44, 71, 117–123), but lacked a principled framework to control false positives rates. In contrast, Bayesian model averaging simultaneously controls the Bayesian FDR and the frequentist FWER, at a level that can be quantified using Doublethink (18). It naturally incorporates uncertainty in which variables to include, which in high-correlation biobank settings can strongly influence the evidence of direct effects. In this study it allowed us to test null hypotheses concerning arbitrary groups of variables, which brought multiple advantages by (i) avoiding the need to manually remove correlated variables pre-analysis, (ii) improving power by combining signals and reducing the stringency of significance thresholds, (iii) conferring flexibility to pursue significant signals *post-hoc* (124), thereby challenging existing notions of fishing for significance, data dredging and *p*-hacking (125).

Despite its strengths, our approach has limitations. We tested only for direct effects, not total effects. This is an important distinction: in many applications, it is necessary to estimate the total effect to understand the likely impact of an intervention on the outcome. The direct effect can differ in magnitude and direction to the total effect, and confusing the two is a pitfall known as the Table 2 fallacy (8). Our Monte Carlo Markov Chain approach was computationally expensive, requiring 3500 CPU hours. Its feasibility depended on asymptotic approximations and a computationally expedient prior (18). These demands limited our ability to model important phenomena like interactions between variables, non-linear effects such as time-since-exposure, or variables with sparse representation like medication use and occupation. Like other methods, Doublethink is subject to *p*-value inflation when testing subsets of highly correlated variables; this can be mitigated by testing groups of correlated variables together (18).

By advancing a more powerful and agnostic approach to identifying direct non-genetic risk factors, our approach has the potential to help advance scientific discovery and bring together the advantages of Bayesian and classical hypothesis testing in biobank-scale settings.

## Data Availability

To access UK Biobank data, researchers must register and submit a research application (https://www.ukbiobank.ac.uk/register-apply). Registration is open to all bona fide researchers for all types of health-related research that is in the public interest. The registration and application process ensures researchers and projects meet UK Biobank's obligations to its participants and funders.

## Acknowledgements

This research has been conducted using the UK Biobank Resource under Application Number 53100. We thank all UK Biobank participants, staff and contributors. We wish to thank Naomi Allen, Jeff Chen, David Eyre, Steven Lin, Gil McVean, Tim Peto and Sarah Walker for comments and advice, and the Mathematisches Forschungsinstitut Oberwolfach, organizers and participants of workshop 2308 *Design and Analysis of Infectious Disease Studies*.

## Supplementary Material

**Table S1** Literature review of published papers analysing COVID-19 outcomes in UK Biobank, containing fields from Web of Science and curated with information on the type of analysis, exclusion criteria, and outcomes and exposures included in the abstract and analysis.

**Table S2** All UK Biobank fields included in the analysis, annotated by field or ICD-10 code, UK Biobank or ICD-10 description and (if a factor) level.

**Table S3** Synonyms and categories of variables used in the interpretation of the literature review of published papers analysing COVID-19 outcomes in UK Biobank.

**Table S4** Categories applied to results in Table 1, prior groupings, for comparison to the literature review

**Table S5** Categories applied to results in Table 2, post hoc groupings, for comparison to the literature review

